# Impact of Daily Weather on COVID-19 outbreak in India

**DOI:** 10.1101/2020.06.15.20131490

**Authors:** Amitesh Gupta, Biswajeet Pradhan

## Abstract

The COVID-19 pandemic has outspread obstreperously in India. As of June 04, 2020, more than 2 lakh cases have been confirmed with a death rate of 2.81%. It has been noticed that, out of each 1000 tests, 53 result positively infected. In order to investigate the impact of weather conditions on daily transmission occurring in India, daily data of Maximum (*T*_*Max*_), Minimum (*T*_*Min*_), Mean (*T*_*Mean*_) and Dew Point Temperature (*T*_*Dew*_), Diurnal Temperature range (*T*_*Range*_), Average Relative Humidity, Range in Relative Humidity, and Wind Speed (*WS*) over 9 most affected cities are analysed in several time frames: weather of that day, 7, 10, 12, 14, 16 days before transmission. Spearman’s rank correlation (r) shows significant but low correlation with most of the weather parameters, however, comparatively better association exists on 14 days lag. Diurnal range in Temperature and Relative Humidity shows non-significant correlation. Analysis shows, COVID-19 cases likely to be increased with increasing air temperature, however role of humidity is not clear. Among weather parameters, Minimum Temperature was relatively better correlate than other. 80% of the total confirmed cases were registered when *T*_*Max*_, *T*_*Mean*_, *T*_*Min*_, *T*_*Range*_, *T*_*Dew*_, and *WS* on 12-16 days ago vary within a range of 33.6-41.3° C, 29.8-36.5° C, 24.8-30.4° C, 7.5-15.2° C, 18.7-23.6° C, and 4.2-5.75 m/s respectively, hence, it gives an idea of susceptible weather conditions for such transmission in India. Using Support Vector Machine based regression, the daily cases are profoundly estimated with more than 80% accuracy, which indicate that coronavirus transmission can’t be well linearly correlated with any single weather parameters, rather multivariate non-linear approach must be employed. Accounting lag of 12-16 days, the association found to be excellent, thus depict that there is an incubation period of 14 ± 02 days for coronavirus transmission in Indian scenario.

## 1. Introduction

In human history, it is apparent that pathogens have caused devastating consequences in social wellbeing and economy (Briz-Redón and Serrano-Aroca, 2020). The recent novel coronavirus disease (COVID-19) is one of the prominent example of such a disastrous event that has grasped the world. The earliest outbreak of COVID-19 caused by Severe Acute Respiratory Syndrome CoronaVirus-2 (SARS-CoV-2) happened in Wuhan, Hubei Province, China during the late December, 2019, (Guan et al., 2020b; Wu and McGoogan, 2020; Zhu et al., 2020; Zu et al., 2020). Because of human-to-human transmissibility of the virus (Wang et al., 2020a; 2020b), the circumstances become progressively unpredictable and vulnerable in terms of transmission of this disease. Considering the rapid turnaround, the World Health Organization (WHO) declared an international public health emergency on January 30, 2020, and later on March 11, 2020, WHO declared this disease as global pandemic due to speedy blowout of infections. Till June 04, 2020, a total of 6,709,724 cases have been affirmed with 5.85% deaths worldwide (https://www.worldometers.info/coronavirus). Despite the fact India has registered its first case on January 29, 2020, the outbreak occurred March 2, 2020 onwards and as of June 04, 2020, a total of 226,722 cases have been confirmed; however, the death rate (2.81%) is quite lower than the worldwide situation.

Clinical investigations on COVID-19 identified respiratory droplets as the most common agent of this infection (Ge et al., 2013; Huang et al., 2020). The reported symptoms are also quite analogous to the other coronavirus diseases such as MERS and SARS, e.g. moderate to high fever with dry cough, and difficulty in breathing attributable to respiratory disorder in early stage, while it causes kidney failure, pneumonia in severe phase (Holshue et al., 2020; Perlman, 2020; Tan et al., 2005; Wang et al., 2020c).

Environmental factors, such as daily weather and long term climatic conditions may affect the epidemiological dynamics of this type of infectious disease (Dalziel et al., 2018; Yuan et al., 2006). Daily air temperature and relative humidity may impact on the transmissions of coronavirus by affecting the persistence of the viral infections within its transmission routes (Casanova et al., 2010). A few studies accounting climate and weather conditions found that these factors considerably affect the spatial distribution along with its incubation period (Bedford et al., 2015; Lemaitre et al., 2019; Sooryanarain and Elankumaran, 2015). At the earliest, Bull (1980) reported that the mortality rate of pneumonia is profoundly associated with the changes in weather condition. Studies have revealed that among different climatic variables the air temperature affects the influenza epidemics mostly in tropical regions (Tamerius et al., 2013) whereas the mid-latitudinal temperate regions experience the influenza diseases epidemics mostly during winter months (Bedford et al., 2015; Sooryanarain and Elankumaran, 2015). Nevertheless, the response to weather pattern on COVID-19 transmission found quite debatable, since, the studies carried out in different countries in the world suggested an existing correlation between weather and COVID-19 pandemic likewise that it occurs with other influenza infections (Ficetola and Rubolini, 2020; Liu et al., 2020; Ma et al., 2020; Oliveiros et al., 2020; Qi et al., 2020; Tosepu et al., 2020). Contradictorily, few studies have reported that meteorological observations are not correlated with outbreak pattern (Jamil et al., 2020; Mollalo et al., 2020; Shi et al., 2020; Xie and Zhu, 2020). Studies carried out by Wang et al., 2020a; Wang et al., 2020b suggested that the spread of disease supposed to be decreased with an increase in temperature. Gupta et al. (2020a) also predicted lowering of transmission in warmer conditions in India. However, in view of the long term climate record, Gupta et al., 2020b found, comparatively hot areas in India are possibly going to be more affected by this disease. Besides, the incubation period of COVID-19 also may vary spatially. The WHO reported an incubation period of 2-10 days for COVID-19 based on worldwide observation (Novel Coronavirus(2019-nCoV) Situation Report - 7, 2020) while the National Health Commission in China had initially estimated an incubation period of 10-14 days for China (https://www.aljazeera.com/news/2020/01/chinas-national-health-commission-news-conference-coronavirus-200126105935024.html) and the Centres for Disease Control and Prevention in United States of America estimate this incubation period of 2-14 days (https://www.cdc.gov/coronavirus/2019-ncov/symptoms-testing/symptoms.html). On other hand, Bai et al., 2020; Guan et al., 2020a reported incubation period of around 20days. Since no such study has investigated the impact of daily weather on COVID-19 transmission in Indian context as well as the incubation period of this disease in India is not mentioned anywhere to date, there is a need of comprehensive study for Indian scenario. Thus, the present study is aimed to understand the trends, abrupt changes and influence of daily weather conditions in COVID-19 transmission in India. We have also investigated the incubation period of this disease based on five timeframes, specifically on the day of the case, within 7, 10, 12, 14, and 16 days of the case.

## 2. Data and Methodology

### 2.1 Data collection

India, the largest country in South Asia, extended from 6° N to 38° N and 68° E to 98° E, comprising a land area of 3.287 million sq. km. with a total population of more than 1.2 billion (Census, 2011). The data of daily COVID-19 cases were collected from the official website of the Ministry of Health of India (https://www.mohfw.gov.in). Among 725 districts in India, more than 85% has reported multiple confirmed cases. Several studies have reported that the disease spread at a higher rate in the cities where population is very high (Ahmadi et al., 2020; Bonasera and Zhang, 2020; Casanova et al., 2010; Kang et al., 2020; Rocklöv and Sjödin, 2020). Thus, among 53 ‘million cities’ (where the total population is more than one million) in India, 9 cities have been selected for this study (Fig. 1), from where more than 79% of total cases in India have been reported till June 4, 2020. The trend of confirmed cases over those cities along with comparison of trend of daily transmission in entire country is highly increasing (Fig. 2). The weather data were collected from https://www.wunderground.com. Fig. 3 shows the prevailing weather conditions in terms of Maximum, Minimum and Mean Temperature, Diurnal Range in Temperature, Dew Point Temperature, Average Relative Humidity, Diurnal Range in Relative Humidity and Wind Speed in those cities. It exhibits that there were variations in weather conditions in different cities, hence, this study will signify how spatially varying weather conditions influence the pattern of COVID-19 transmission in India.

**Fig. 1.**
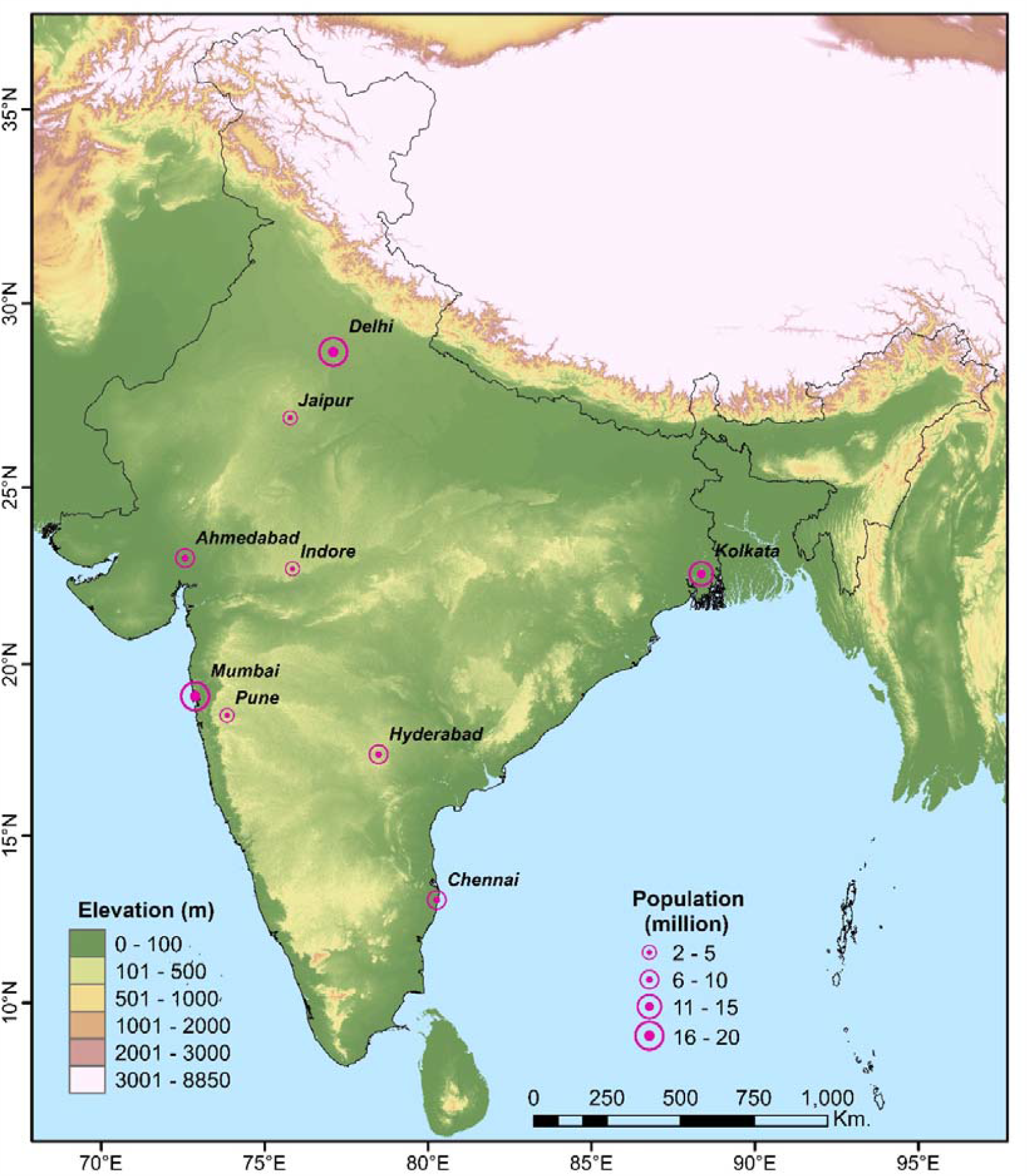
Location of the selected cities in India along with the total population of those cities.

**Fig. 2.**
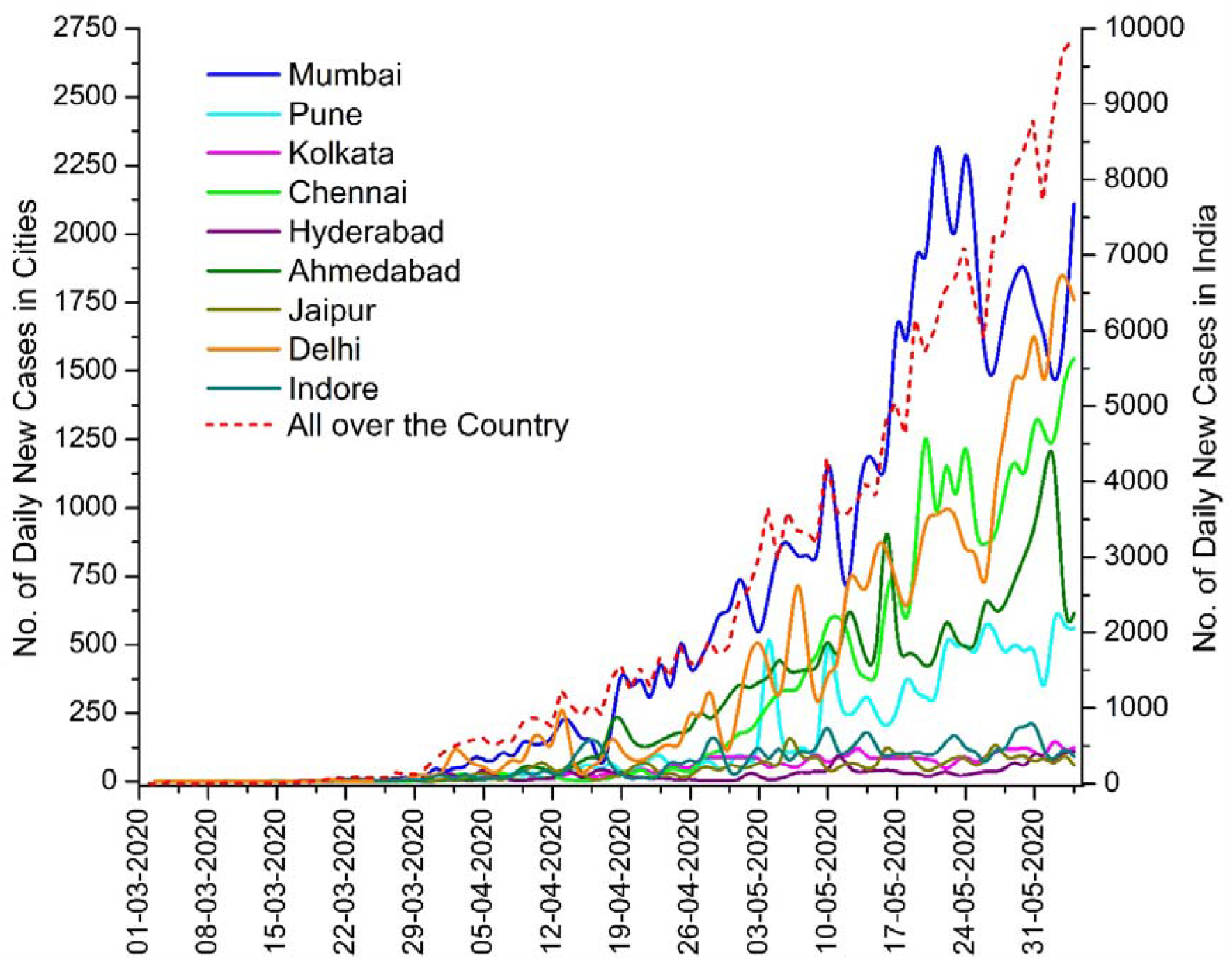
Trend of daily confirmed cases over selected cities and all-over the country.

**Fig. 3.**
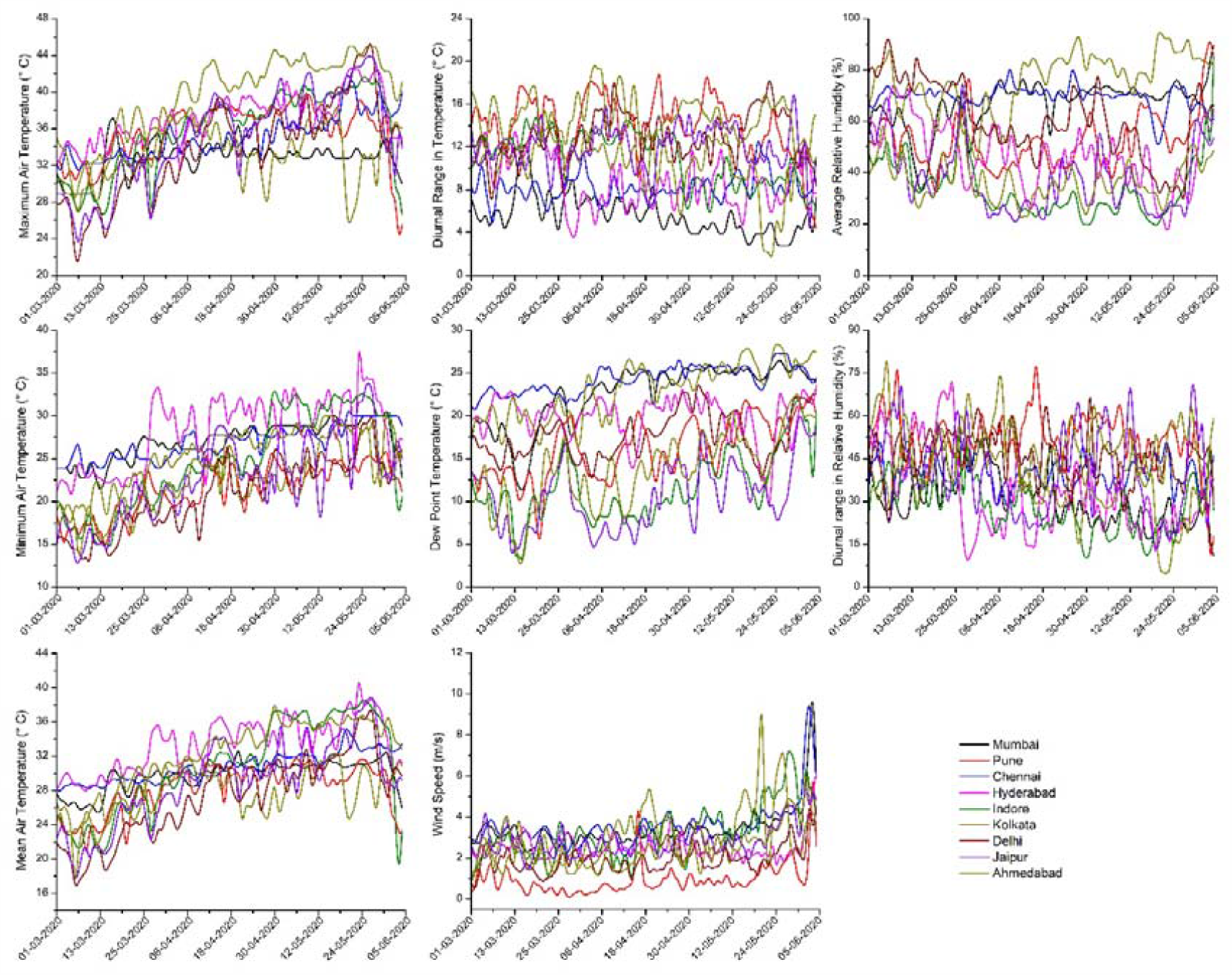
Pattern of Daily Weather over the selected cities in India.

### 2.2 Spearman’s correlation test

Spearman’s rank correlation coefficient (*r*_*s*_) is implemented to define the association between a number of daily new cases and weather parameters. It summarizes how well the association between daily transmission and weather parameters can be demarcated. The coefficient can be calculated via the following equation –

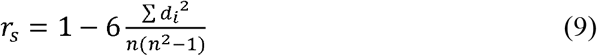

where, *n* represents the number of alternatives, and *d*_*i*_ is the difference between the ranks of two parameters.

### 2.3 Support Vector Machine

Support Vector Machine (SVM) is an extensively utilized machine learning technique. It is performed on the basis of statistical auto-adaptation and structural risk minimization principle (Tien Bui et al., 2012). By creating hyper-plane, the nonlinearity in the input dataset is reshaped into the linearity (Jebur et al., 2014). Here, kernel function is the key factor behind this data transformation. Using the assigned training dataset, SVM put the original input into a higher dimensional feature space, then finds the supreme fringe of separation among the observations and constructs a hyper-plane at the centre of that extreme margin (Marjanović et al., 2011). Support vectors are nothing but the nearest training points to the produced hyper plane. Thus, this model adapt itself by input observations and create hyper-plane and identify the support vectors and thereafter acted on the input variables of testing dataset to estimate the predicted variable. Further insights about the mathematical computations and procedures work in SVM can be found in several literature such as Pradhan, 2013; Tehrany et al., 2015, 2014; Tien Bui et al., 2012. However, the accuracy of estimation depends on the kernel type selected during the training of the model (Yao et al., 2008). The Radial basis function (RBF) kernel produce preferred exactness than linear, polynomial and sigmoid kernels due to its higher capability in interpolation (Song et al., 2011).

Early observation by Gupta et al. (2020c) noted that transmission in India is likely to be higher over those area which are located in lower altitudes and having higher population. Thus, we also incorporate elevation and population of those selected cities along with the daily weather and estimate the log-transformed value of daily COVID-19 cases (Eq. 10).

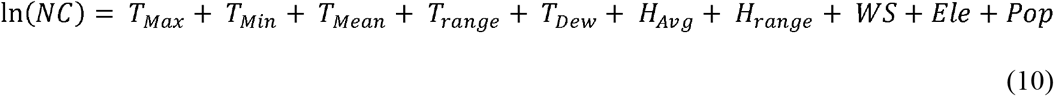

where, *NC* is the number of New Confirmed Case, *T*_*Max*_ is Maximum Air Temperature (° C), *T*_*Min*_ is Minimum Air Temperature (° C), *T*_*Mean*_ is Mean Air Temperature (° C), *T*_*range*_ is Temperature Range (° C), *T*_*Dew*_ is Dew point Temperature (° C), *H*_*Avg*_ is Average Relative Humidity (%), *H*_*Avg*_ is Range of Relative Humidity (%), *WS* is Wind Speed, *Ele* is Elevation (m), *Pop* is total Population.

70% of the total observation was used as training dataset and rest used for testing. The accuracy of estimation was evaluated in terms of R^2^, Root Mean Square Error (RMSE) and Mean Bias (MB).

## 3. Results and discussion

The Spearman’s correlation analysis (Table 1) shows that there were mostly significant but considerably low correlation lies between the number of daily new case and weather condition. Among weather parameters, only *T*_*range*_ and *H*_*range*_ are negatively correlated with the daily transmission, however, correlation for *T*_*range*_ is non-significant in all time span. Hence, the diurnal range of temperature is not significantly associated with COVID-19 transmission in India. *H*_*avg*_ is significant on the day of transmission up to 10 days ago of transmission, while *H*_*range*_ is significant 12-16 days ago of transmission; it suggest that the role of humidity is quite complex and needed to be investigated further in depth. On other hand, the analysis indicate that the *T*_*max*_, *T*_*min*_, *T*_*mean*_, *T*_*Dew*_ on the day of the transmission has the lowest correlation and it improves at its best with a time lag of 14 days. In other words, the maximum, minimum, mean and dew point temperature on 14 days ago of transmission is closely related with a number of infections. Interestingly, *T*_*min*_ is found better related than *T*_*mean*_ *T*_*max*_ *T*_*Dew*_. Therefore, places with higher minimum temperature are more susceptible for COVID-19 transmission in India. WS also found to be positively correlated with daily transmission, which may infer that virus might be able to transmigrate with high wind. Since, most of the weather parameters including WS, are better correlate with the daily confirmed cases with a time lag of 14 days, it indicate an approximate incubation period of around 14 days for this disease in the Indian scenario.

**Table 1.** Result of Spearman’s correlation test

|  | On that | 7 Days | 10 Days | 12 Days | 14 Days | 16 Days |
| --- | --- | --- | --- | --- | --- | --- |
| Parameters | Day | ago | ago | ago | ago | ago |
| Maximum Temperature | 0.161* | 0.198* | 0.231* | 0.336* | 0.347* | 0.272* |
| Minimum Temperature | 0.244* | 0.285* | 0.319* | 0.417* | 0.436* | 0.351* |
| Mean Temperature | 0.199* | 0.248* | 0.287* | 0.409* | 0.430* | 0.337* |
| Temperature Range | -0.032 | -0.043 | -0.066 | -0.064 | -0.075 | -0.054 |
| Dew Point Temperature | 0.222* | 0.235* | 0.261* | 0.238* | 0.269* | 0.238* |
| Average Relative Humidity | 0.128* | 0.1* | 0.09* | 0.002 | 0.008 | 0.034 |
| Humidity Range | -0.016 | -0.024 | -0.056 | -0.13* | -0.149* | -0.095* |
| Wind Speed | 0.108* | 0.133* | 0.163* | 0.221* | 0.255* | 0.193* |

\*Significant at 0.05 significance level.

Fig. 4 shows the validation of estimated daily confirmed cases for all time spans using non-linear multivariate Support Vector Regression Model with RBF kernel. The model performance in terms of R^2^, RMSE, MB are represented in Table 2. It depicts that SVM based regression model is very efficient to establish the complex relationship among different weather parameters with the daily transmission of COVID-19, however, it exhibits an underestimation for very high values (>1200 cases). Hence, it make us understood that any single weather parameter is not enough to linearly correlate the daily transmission, rather than that, the non-linear multivariate approach is efficient to estimate the daily transmission in India with high accuracy. Correlation analysis has evidently stipulated a relatively higher degree of association for daily new cases with most of the parameters only when the time lag of 14 days is taken in to consideration. The SVM based regression model also performs remarkably well with a time lag of more than 12 days. These together suggest a conspicuous incubation period of 12-16 (14 ± 02) days for this transmission in India. In order to better understand the influence of varying weather conditions, the response curve of significant parameters to cumulative confirmed cases was framed (Fig. 5), which reveal that there is an acute range in weather parameters for which the transmission is highly susceptible. 80% of the total confirmed cases were registered when *T*_*Max*_, *T*_*Mean*_, *T*_*Min*_, *T*_*Range*_, *T*_*Dew*_, and *WS* on 12-16 days ago vary within a range of 33.6-41.3° C, 29.8-36.5° C, 24.8-30.4° C, 7.5-15.2° C, 18.7-23.6° C, and 4.2-5.75 m/s respectively. Hence, it gives an idea of susceptible weather conditions for such transmission in India. In other words, the areas experiencing such weather pattern in India must have been affected by this disease.

**Table 2.** Result of Validation of SVM based regression for estimating daily transmission.

|  | R<sup>2</sup> | RMSE | MB |
| --- | --- | --- | --- |
| On that Day | 0.6414 | 199.2929 | -48.1804 |
| 7 Days ago | 0.7015 | 202.1743 | -40.0377 |
| 10 Days ago | 0.8286 | 223.1949 | -42.0560 |
| 12 Days ago | 0.8503 | 186.0126 | -66.9880 |
| 14 Days ago | 0.8680 | 178.3891 | -43.6459 |
| 16 Days ago | 0.8714 | 202.2428 | -60.0658 |

**Fig. 4.**
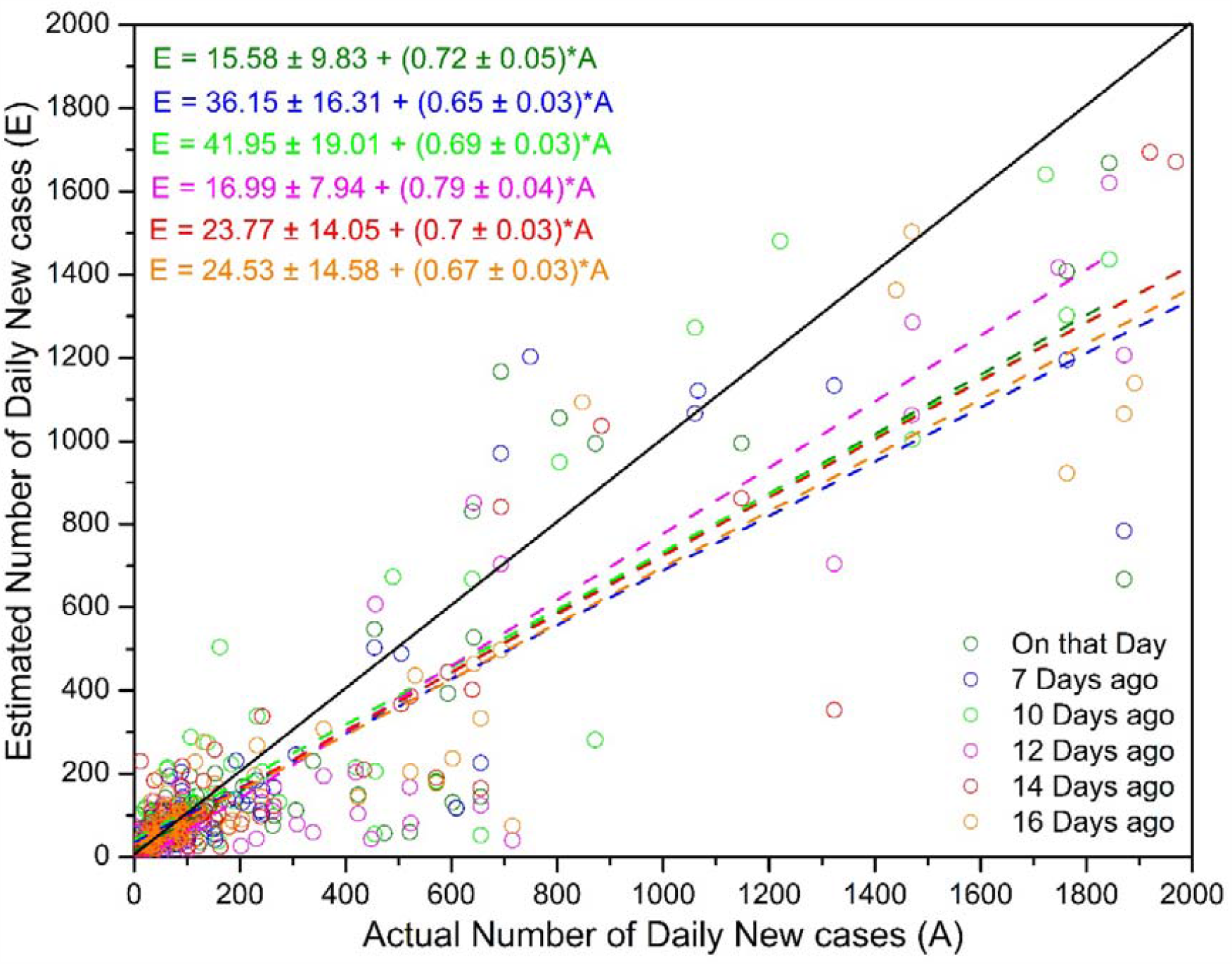
Validation of SVM based regression model for estimating daily transmission.

**Fig. 5.**
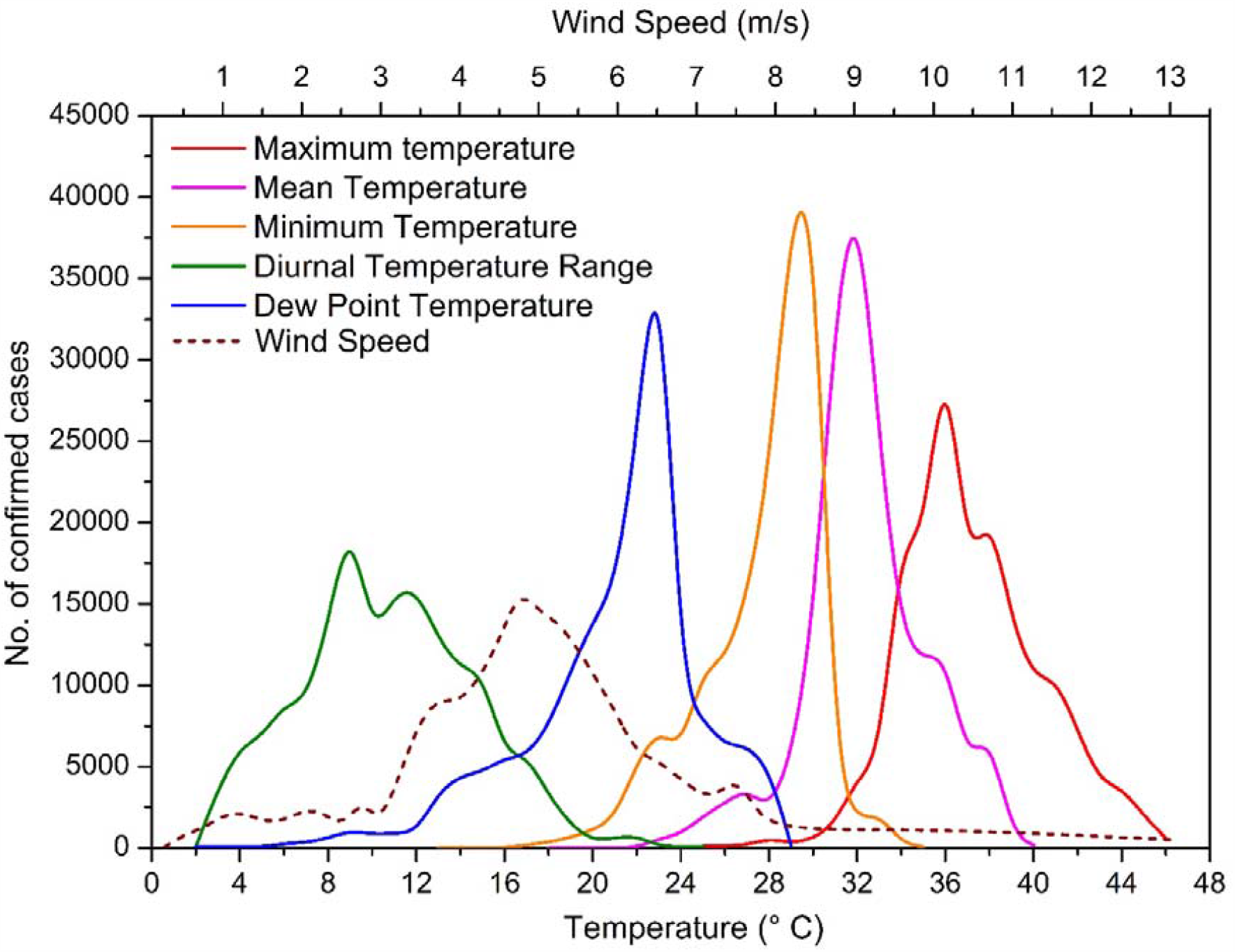
Influence of weather parameters on count of confirmed cases with a lag of 12-16 days.

## 4. Conclusion

Unlike most of the studies, the present study investigated the impact of daily maximum, minimum, mean, and dew point temperature, temperature range, average humidity, humidity range and wind speed on that day, as well as within 7, 10, 12, 14, and 16 days of the confirmed cases of COVID-19 in the Indian context. The analysis revealed that the count of confirmed cases significantly correlated with a certain range of weather conditions. Thus, instead of linear correlation, SVM based regression approach efficaciously resolve this complex association and able to estimate daily cases of transmission using the weather inputs. However, the positive correlation between daily transmission and air temperature as well as wind speed designates that the daily transmission in highly populated areas in India has been responsively increased during current summer days. A prominent incubation period of 14 ± 02 days has also been identified, which was a little higher than what WHO had prescribed early in March. Therefore, in the prevailing weather conditions in India, the SARS-CoV-2 can be disseminated into the surrounding environment for around two weeks after being grieved from any other contaminant. This study had faced several limitations since many other major affected cities were not able to incorporate due to lack of data availability. Besides, the count of immigrants from abroad or other cities and have been quarantined were not available; these might can enhance the exactitude of the current analysis.

## Data Availability

Data of daily confirmed cases available from the website of Health Ministry, Govt. of India. Weather data acquired from https://www.wunderground.com.

## CRediT authorship contribution statement

**Amitesh Gupta**: Conceptualization, Methodology, Investigation, Visualization, Writing – original draft. **Biswajeet Pradhan**: Writing – review and editing, Supervision.

## Acknowledgement

This research had not any funding support. Mr. Sumit Das is highly acknowledged for his continuous supports and substantial editing to improve the quality of paper and proof reading. Dr. Arkadeb Ghosh is also acknowledged for his kind help during the work.

